# Olfactory Training: Recommendation Frequency Amongst Rhinologists

**DOI:** 10.1101/2024.10.31.24315756

**Authors:** Sherina R. Thomas, Vincent L. Nguyenkhoa, Jose L. Mattos, Steven D. Munger

## Abstract

Despite the prevalence and health impact of olfactory dysfunction (OD), therapeutic options remain limited. One potential therapy is olfactory training (OT), where patients repeatedly sniff a small set of odors over several weeks with the goal of improving olfactory function. Although double-blinded, placebo-controlled clinical trials supporting the efficacy of OT are lacking, anecdotal evidence suggests that the procedure is widely recommended to patients with anosmia or other olfactory disorders. To better understand the adoption of OT in clinical practice, we recruited a convenience sample of 95 rhinologists and other otolaryngologists to complete an online survey that assessed how often, and under what conditions, they recommend OT to their patients with OD. Our survey revealed that the majority of responding otolaryngologists (93.7%) routinely recommend OT. Further, they recommend OT more often for quantitative than for qualitative disorders, and most frequently for post-viral and idiopathic OD etiologies. These findings support the perception that OT is commonly recommended in otolaryngological practice for the treatment of OD.

**Key Points:**

- Most surveyed otolaryngologists routinely recommend olfactory training (OT).
- OT is recommended more often for quantitative than qualitative olfactory dysfunction (OD).
- OT is most often recommended for post-viral and idiopathic OD etiologies.

## Introduction

Olfactory dysfunction (OD) affects ∼22% of adults.^1^ Characterized by reduced, absent, or distorted smell function, OD can negatively impact safety, diet, social relationships, mental health, and even lifespan.^2^ Despite the prevalence of OD and its significant health burden, few effective treatments are available, and even those options may only be appropriate for certain patient populations.^3^

One potential therapy is olfactory training (OT),^4^ which involves focused sniffing of multiple odors twice daily for 3-6 months.^4,5^ Numerous studies support the ability of OT to improve olfactory function in anosmic and hyposmic patients.^2,3^ However, large, blinded studies remain necessary to clearly differentiate OT-dependent improvements in smell function from spontaneous recovery.^3^ Compliance is challenging for patients,^6^ in part because of the lengthy commitment with uncertain benefit. Anecdote suggests that despite these challenges, OT is widely recommended by providers to patients with OD. To furnish a more rigorous estimate, we recruited a convenience sample for a survey-based study to determine how often, and under what conditions, otolaryngologists recommend OT to their patients with OD.

## Methods

This study was approved by the University of Virginia (UVA) Institutional Review Board. We distributed two invitations for an anonymous, 13-item survey (**Supporting Information**) to 844 members of the American Rhinologic Society (ARS) via email listserv. Interested participants were directed to a UVA REDCap server to complete a multiple-choice survey, with some questions offering text boxes for expanded responses. Questions assessed provider demographics including subspeciality and practice setting, OD diagnostic regimen, and OT recommendation habits. We received ninety-five responses over four weeks, yielding an 11.3% response rate.

Results are reported as raw numbers and percentages. Preliminary analyses prompted the assessment of two potential associations using chi-square tests (d.f. = 1, p=0.05). The first was to assess whether the use of psychophysical smell tests for OD diagnosis varied between otolaryngologists based on practice setting (private vs. hospital/academic) or subspeciality (rhinologists vs. general otolaryngologists). The second was to assess whether these same subsets differed in their likelihood to recommend OT to their patients with OD.

## Results

Of all respondents, 27.4% (n=26) self-identified as general otolaryngologists, 71.6% (n=68) as rhinologists, and 1.1% (n=1) as another subspecialist (see **Supporting Information, Table 1** for all multiple choice and text box responses). The survey found that 60.0% (n=57) of respondents practiced in an academic medical center, 13.7% (n=13) in non-academic or community hospitals, and 24.2% (n=23) in private practice. All respondents saw patients with complaints of smell dysfunction.

A subset of respondents (60.0%; n= 57) used psychophysical smell tests in their diagnostic regimen for OD, with 77.2% (n=44) routinely using the University of Pennsylvania Smell Identification Test (UPSIT),^7^ 17.5% (n=10) using Sniffin’ Sticks,^8^ and 7.0% (n=4) using other tests (**Supporting Information**). Respondents in academic medical centers or non-academic hospitals were more likely to employ psychophysical smell tests than those in private practice (X^2^ = 11.05, p = 0.0009). However, there was no difference in psychophysical test usage between rhinologists and other otolaryngologists (X^2^ = 2.21; P = 0.14).

Respondents overwhelmingly reported recommending OT (93.7%; n=89) for patients with OD (**Figure 1A**), with 56.2% (n=50) recommending it to >75% of their patients. The likelihood of recommending OT did not differ with subspeciality (X^2^ = 1.47; p = 0.23) or practice type (X^2^ = 0.20; p = 0.66). OT is routinely recommended for all types of OD, though most commonly for quantitative impairments (anosmia and hyposmia) (**Figure 1B**). Further, while OT is most often recommended to patients with post-viral or idiopathic OD, many providers recommend OT regardless of etiology (**Figure 1C**). Those who recommended OT reported confidence in current research backing OT’s efficacy (52.8%, n=47), personal experience (44.9%, n=40), or minimal treatment risks (47.2%, n=42). 66.7% (n=4) of those who did not recommend OT noted a lack of convincing research supporting its use. The majority (55.1%, n=49) of respondents recommend OT to patients under 18 years of age. Finally, 51.7% (n=46) of respondents reported that their recommendation of OT is not impacted by the duration of a patient’s smell loss.

**Figure 1.**
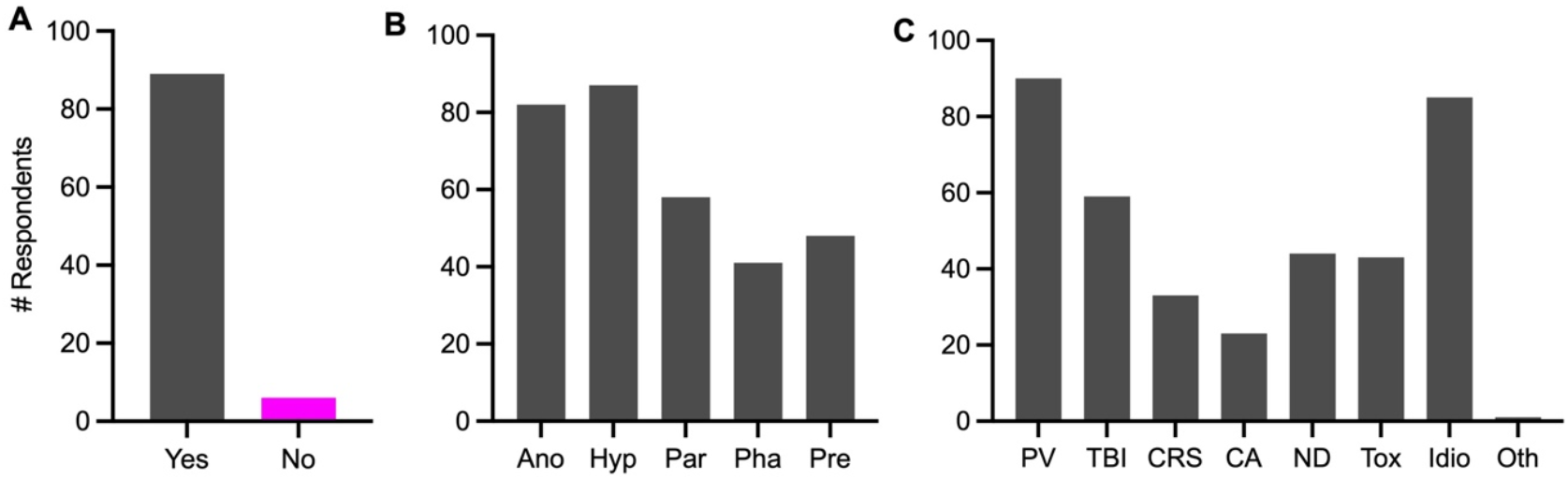
Responses to select survey questions about smell training. **(A)** Those who do (grey) or do not (magenta) recommend smell training to their olfactory disorders patients. N=95 **(B)** Olfactory disorders for which respondents recommend smell training (respondents could choose more than one). Ano, anosmia; Hyp, hyposmia; Par, parosmia; Pha, phantosmia; Pre, presbyosmia. N=89 **(C)** Etiologies for which respondents recommend smell training (respondents could choose more than one). PV, post-viral; TBI, traumatic brain injury; CRS, chronic rhinosinusitis; AR, allergic rhinitis; ND, neurodegenerative disease; Env, environmental exposures (e.g., toxins); Idio, idiopathic. N=89

## Discussion

The efficacy of OT remains elusive due to mixed evidence regarding its benefit for the broad range of OD etiologies.^3^ Despite this, the vast majority of surveyed otolaryngologists routinely recommend OT for adult patients with OD. The lower frequency of recommending OT to pediatric patients could reflect a lower adherence to treatment guidelines in adolescent populations, but this remains unassessed. Findings further suggest that OT is most recommended for patients with post-viral or idiopathic anosmia and hyposmia, likely reflecting the existing research that has focused on these populations. ^2^ Even so, it appears that clinical usage is outpacing clinical evidence, as many respondents recommended OT regardless of patient presentation.

Hospital-based providers were more likely to employ psychophysical tests for OD diagnosis than those in non-hospital private practice, perhaps reflecting cost considerations, protocol flexibility, or resource availability.^9^ However, OD diagnostic regimens and OT recommendation habits did not significantly differ between rhinologists and general otolaryngologists, suggesting that specialized rhinology training does not greatly influence these practices.

### Limitations and Future Directions

This convenience sample, recruited through the ARS listserv, focused on providers with rhinological interests, which likely enriched the sample for those familiar with OT. Future assessments that increase the participation of general otolaryngologists, non-otolaryngologists, and international practitioners would complement the findings reported here. Nevertheless, the high frequency of OT recommendation by respondents supports the conclusion that OT is commonly recommended in otolaryngological practice for the treatment of OD.

## Data Availability

All data produced in the present work are contained in the manuscript.

