## Supporting Information for "Olfactory Training: Recommendation Frequency Amongst Rhinologists"

### Smell Training Recommendation Survey

**Please complete the survey below.**

**Objective**
To determine the frequency with which rhinologists recommend smell training to patients with olfactory disorders.

**Definitions**
Olfactory (smell) disorders include the following:
*Hyposmia (aka microsmia):* a reduced ability to perceive smells
*Anosmia:* the complete inability to perceive smells
*Parosmia:* distorted smell perceptions
*Phantosmia:* phantom smell perceptions
*Presbyosmia*: age-related smell loss

Smell training (also known as olfactory training or smell re-training) is the daily, intentional smelling of 1-4 odors over a period of weeks with the goal of promoting the recovery of normal smell function.

**Survey questions**

1. Which best describes your medical specialty?
    A. General otolaryngology
    B. Rhinology
    C. Other otolaryngology subspecialty
    D. Other (text box)
2. Is your medical practice in:
    A. An academic medical center
    B. A non-academic or community hospital
    C. A non-hospital private practice
    D. Other (text box)
3. Do you see patients with complaints of smell or taste dysfunction?
    A. Yes
    B. No [If No, terminate]
4. Do you include psychophysical smell tests (such as the University of Pennsylvania Smell Identification Test (UPSIT) or Sniffin' Sticks) as part of your diagnostic regimen for patients with a complaint of smell or taste dysfunction?
    A. Yes
    B. No [If No, skip to Q6]

5. Which smell test do you typically employ?
    A. UPSIT
    B. Sniffin' Sticks
    C. Other (text box)
6. For those patients where you diagnose a smell disorder, do you recommend smell training as a therapy?
    A. Yes [If Yes, skip to Q8]
    B. No
7. Why do you NOT recommend smell training? (Choose all that apply)
    A. I was unaware of it
    B. Published studies have not convinced me it is effective
    C. In my experience I have not found it to be useful
    D. My patients do not adhere to the regimen
    E. Other (text box)

    [If Q7 answered, terminate]
8. Why do you recommend smell training? (Choose all that apply)
    A. In my experience it helps at least some of my patients with smell disorders
    B. I am convinced by the scientific and clinical literature that it is an effective therapy for at least some patients with smell disorders
    C. While I am unsure of smell training's effectiveness as a therapy for smell disorders, I see no negative side effects or potential harms of my patients trying it
    D. Other (text box)
9. For which olfactory disorder(s) do you recommend smell training? (Choose all that apply)
    A. Anosmia
    B. Hyposmia
    C. Parosmia
    D. Phantosmia
    E. Presbyosmia
10. For which olfactory dysfunction etiologies do you recommend smell training? (Choose all that apply)
    A. Post-viral
    B. Traumatic brain injury or head trauma
    C. Chronic rhinosinusitis
    D. Chronic allergies
    E. Neurodegenerative disease
    F. Toxic chemical exposure
    G. Idiopathic
    H. Other (text box)

11. For which age group do you recommend smell training? (Choose all that apply)
    A. Under 18 years old
    B. 18-40 years old
    C. 41-65 years old
    D. Over 65 years old
12. I recommend smell training:
    A. Up to 6 months after the onset of the disorder
    B. Up to 1 year after the onset of the disorder
    C. Up to 2 years after the onset of the disorder
    D. The duration of the smell loss does not impact my recommendation
    E. Other (text box)
13. I recommend smell training to approximately ______ of my smell disorders patients.
    A. <25%
    B. 25%-49%
    C. 50-75%
    D. >75%

Thank you for your participation in this survey. Have a nice day!

**Table 1:** Smell Training Survey Response Numbers

| *Question Number* | *Answer Choice* | *# of Respondents* |
|---|---|---|
| *1* | A | 26 |
| | B | 68 |
| | C | 1 |
| | D | 0 |
| *2* | A | 57 |
| | B | 13 |
| | C | 23 |
| | D | 2 |
| *3* | A | 95 |
| | B | 0 |
| *4* | A | 57 |
| | B | 38 |
| *5** | A | 43 |
| | B | 8 |
| | C | 5 |
| *6* | A | 89 |
| | B | 6 |
| *7* | A | 1 |
| | B | 4 |
| | C | 2 |
| | D | 0 |
| | E | 1 |
| *8* | A | 40 |
| | B | 47 |
| | C | 42 |
| | D | 0 |
| *9* | A | 82 |
| | B | 87 |
| | C | 58 |
| | D | 41 |
| | E | 48 |
| *10* | A | 89 |
| | B | 58 |
| | C | 32 |
| | D | 22 |
| | E | 43 |
| | F | 42 |
| | G | 83 |
| | H | 3 |
| *11* | A | 49 |
| | B | 87 |
| | C | 87 |
| | D | 80 |
| *12* | A | 13 |
| | B | 19 |
| | C | 11 |
| | D | 46 |
| | E | 0 |
| *13* | A | 8 |
| | B | 10 |
| | C | 21 |
| | D | 50 |

*Number of responses for each survey answer choice, sorted by question. *One respondent chose not to answer Q5, a question nested under Q4.*

**Table 2:** Smell Training Survey Textbox Responses

| Question | Response |
|---|---|
| Is your medical practice in:<br>E. An academic medical center<br>F. A non-academic or community hospital<br>G. A non-hospital private practice<br>H. Other (text box) | "Affiliated professor [named university redacted]" |
| | "Private hospital" |
| Which smell test do you typically employ?<br>A. UPSIT<br>B. Sniffin' Sticks<br>C. Other (text box) | "Connecticut" |
| | "NIH" |
| | "BSIT/Sniff sticks" |
| | "I use both. Sniffin sticks prior to any experimental therapy. UPSIT for routine assessment." |
| | "Connecticut Chemosensory Clinical Research Center (CCCRC)" |
| Why do you NOT recommend smell training? (Choose all that apply)<br>A. I was unaware of it<br>B. Published studies have not convinced me it is effective<br>C. In my experience I have not found it to be useful<br>D. My patients do not adhere to the regimen<br>E. Other (text box) | "It is not a choice, but I do use it in rare specific circumstances. Your survey would be better if you listed often, sometimes, rarely, never rather than yes /no." |
| For which olfactory dysfunction etiologies do you recommend smell training? (Choose all that apply)<br>A. Post-viral<br>B. Traumatic brain injury or head trauma<br>C. Chronic rhinosinusitis<br>D. Chronic allergies<br>E. Neurodegenerative disease<br>F. Toxic chemical exposure<br>G. Idiopathic<br>H. Other (text box) | "Presbyosmia" |
| | "Postoperative skull base" |
| | "Any etiology" |
